# SARS-CoV-2 Load does not Predict Transmissibility in College Students

**DOI:** 10.1101/2021.03.02.21252105

**Authors:** Di Tian, Zhen Lin, Ellie M. Kriner, Dalton J. Esneault, Jonathan Tran, Julia C. DeVoto, Naima Okami, Rachel Greenberg, Sarah Yanofsky, Swarnamala Ratnayaka, Nicholas Tran, Maeghan Livaccari, Marla Lampp, Noel Wang, Scott Tim, Patrick Norton, John Scott, Tony Y. Hu, Robert Garry, Patrice Delafontaine, Lee Hamm, Xiao-Ming Yin

## Abstract

SARS-CoV2 is highly contagious and the global spread has caused significant medical, social and economic impacts. Other than vaccination, effective public health measures, including contact tracing, isolation and quarantine, is critical for deterring viral transmission, preventing infection progression and resuming normal activities. Viral transmission is affected by many factors but the viral load and vitality could be among the most important ones. Although in vitro culture studies have indicated that the amount of virus isolated from infected people determines the successful rate of virus isolation, whether the viral load carried at the individual level would affect the transmissibility was not known. We aimed to determine whether the Ct value, a measurement of viral load by RT-PCR assay, could differentiate the spreader from the non-spreader in a population of college students. Our results indicate that while at the population level the Ct value is lower, suggesting a higher viral load, in the symptomatic spreaders than the asymptomatic non-spreaders, there is a significant overlap in the Ct values between the two groups. Thus Ct values, or the viral load, at the individual level could not predict the transmissibility. Our studies also suggest that a sensitive method to detect the presence of virus is needed to identify asymptomatic persons who may carry a low viral load but can still be infectious.

## Introduction

The rapid spread of SARS-CoV-2 has caused a global pandemic with serious impact on all aspects of human life. Deterrence of viral transmission through public health measures, including contact tracing, isolation and quarantine, is critical for infection control required to resume normal activities. Unlike two other betacoronavirus that had caused previous local epidemics, SARS-CoV and MERS-CoV, SARS-CoV-2 exhibits a distinct replication and transmission kinetics. It replicates more rapidly in the human upper respiratory tract, which helps its transmission through asymptomatic viral carriers and facilitates a fast spread of SARS-CoV-2. The relatively lower fatality rate (CFR or case fatality ratio) of SARS-CoV-2 (2% compared to SARS-CoV’s 10% and MERS-CoV’s 34%) may also contribute to its high transmissibility [1]. SARS-CoV-2 is highly contagious with an estimated reproductive number (Ro) of 3.5 [2], but significant variations exist among individuals with some being super spreaders. This is much higher than the Ro of seasonal flu (1.3) and SARS-CoV (0.86-1.83) [3, 4].

The viral load in an infected person could affect the level of infectivity. Several studies have found that successful isolation of virus from patient samples depended on viral load as measured by the cycle threshold (Ct) value of the RT-PCR assay, which was thus suggested to correlate with infectivity [5-10]. A cutoff Ct value between 32 and 35 was proposed to guide isolation practices [5-10]. However, it was not clear whether the *in vitro* culture results could reflect actual viral spread in persons and whether Ct values could actually be used to guide isolation and quarantine decisions.

The effective way to block the viral transmission is to identify, isolate and treat the infected persons, and to track down and quarantine those having close contact with the infected ones. As the infection involves more and more people, individual communities or regions may be forced to be shut down. All social activities related to work, study and leisure will be significantly affected with tremendous impacts on the economy, the society and the overall personal health condition. It is thus important to understand better the dynamics of viral transmission and examine whether certain surrogate measurement may be used to determine SARS-Cov2 transmissibility. We thus aimed to determine whether viral load, as measured by Ct values, could be used to provide a level of prediction in a population of college students. We compared the viral load of the spreader and that of the non-spreader and found these values were largely overlapped. It is thus not possible to predict viral transmissibility based on Ct values at the individual level.

## Materials and Methods

### Study population

Undergraduate students at the age of < 23 years of old were selected in this retrospective study. These students were participants in the on-campus education activities and were tested multiple times in the period between September 1^st^ 2020 and October 31^st^, 2020. This study included only students who were tested in the Molecular Pathology Laboratory of the Department of Pathology and Laboratory, Tulane University School of Medicine because the Ct values were obtained using the same testing method in the same laboratory for all the included subjects.

### Sample collection, processing and RNA extraction

Nasopharyngeal swab specimens were collected following current CDC guidelines. All samples were stored at 4°C before delivering to the testing laboratory. Upon receiving, samples were inactivated at 60°C for 30min in a forced-air oven (Thermo Fisher Scientific, catalog No. 151030510). RNA was extracted using a KingFisher Flex Magnetic Particle Processor with 96 Deep-Well Head (Thermo Fisher Scientific, catalog No. 5400630) and the MagMax Viral/Pathogen Nucleic Acid Isolation Kit (Thermo Fisher Scientific, catalog No. A48310). An MS2 Phage Control was included as an extraction control in the original sample before total RNA extraction.

### TaqPath ^™^ RT-PCR COVID-19 Combo Kit assay

This is an FDA-approved assay under EUA. Multiplex RT-qPCR was performed according to the manufacturer’s instructions (Thermo Fisher Scientific, catalog No. A47814). Viral nucleic acids were detected using primers and probes targeting the N, S and ORF1ab genes. A pair of primers against the extraction controls (MS2) were also included in the same reaction. RT-qPCR reactions were performed on either an ABI7500 FAST DX Real-Time PCR instrument (Thermo Fisher Scientific, catalog No. 4406985) or a QuantStudio^™^ 5 Real-Time PCR instrument (Thermo Fisher Scientific, catalog No. A34322). Positive samples were identified using the Applied Biosystems^™^ COVID-19 Interpretive Software v1.3 (for ABI7500 FAST DX) or Applied Biosystems^™^ COVID-19 Interpretive Software v2.3 (for QuantStudio^™^ 5).

### Ct values analysis

The Ct values for the three viral genes (N, S, and ORF1ab) and extraction control were determined individually by an analytical software SDS v1.4.1 (for ABI7500 FAST DX) or QuantStudio Design and Analysis Desktop Software v1.5.1 (for QuantStudio^™^ 5). The Ct values reported in this study are the average of the values for the three viral genes. Selection of Ct values of 24 or 32 as a threshold (Supplemental Fig. 1B,D. F) was based on the literature (see the main text).

### Contact tracing and quarantine

Symptomatic information was collected immediately before sample collection and testing. Contact Tracers received all positive results and made phone calls to reach positive cases. They interviewed the positive cases to identify close contacts. In addition, they helped to establish the quarantine procedure. The information of the index cases and the contacted was recorded.

### Statistical analysis

Data are presented as mean±SD (for the age distribution), mean ± SEM, or median ± interquartile (for the Ct values). The statistical significance is assessed by two-sided unpaired *t* test for age distribution, Mann-Whitney U test or one-way ANOVA for Ct values using Prism Software (version 9, Graphpad Software, Inc. San Diego, CA).

## Results

Colleges represent a unique environment with a dense population of primarily young students and strict control of SARS-CoV-2 transmission is critical for their education mission. Tulane University maintained on-campus educational activities in the fall semester of 2020. We established a high throughput SARS-CoV-2 testing program to support the contact tracing, isolation and quarantine efforts needed to actively restrict viral transmission throughout the campus. From September 1^st^ 2020 to October 31^st^, 2020 we performed a total of 61,982 tests of 7,440 undergraduate students under the age of 23 years old and identified 602 unique positive cases (**Tables 1-2**). Compared to all the students, those tested positive for SARS-CoV-2 were slightly younger, reflecting that more freshmen and sophomores were infected. In addition, male and female students had nearly the same proportion of the infected (49.3% vs 50.7%), consistent with a meta-analysis of 90 reports [11]. However, considering that male students accounted for only 37.5% of all the students screened, the male students had a higher infection rate (10.65%) than the female students (6.56 %) in this cohort.

**Table 1.** The gender distribution of the cases.

|  | All cases |  | Positive cases |  | Infection Rate |
| --- | --- | --- | --- | --- | --- |
|  | n | Proportion (%) | n | Proportion (%) |  |
| **All** | 7440 | 100 | 602 | 100 | 8.09 |
| **male** | 2790 | 37.50 | 297 | 49.34 | 10.65 |
| **female** | 4650 | 62.50 | 305 | 50.66 | 6.56 |

Data reflects the number of unique individual undergraduates being tested in the period of 9/1/2020-10/31/2020. Each individual may be tested multiple time during this period, but each unique positive case is counted only once. Proportion of each gender in the population is calculated by dividing the total individual number by male or female individual numbers for all cases or for positive cases. Infection rate is calculated by dividing the all case number by the positive case number in male or female or all individuals.

**Table 2.** The age distribution of the cases.

| **Age (Yr)** | All cases |  | Positive cases |  | **p** |
| --- | --- | --- | --- | --- | --- |
|  | Mean | SD | Mean | SD |  |
| **All** | 20.28 | 1.31 | 19.64 | 1.11 | <0.001 |
| **male** | 20.32 | 1.32 | 19.75 | 1.20 | <0.001 |
| **female** | 20.25 | 1.30 | 19.54 | 1.02 | <0.001 |

Two-sided unpaired *t* tests were conducted between the positive cases and all cases for all genders, male only or female only.

From this cohort of 602 positive individuals, we identified 195 index cases with one or more reported close contacts who were then tested during their mandated 14-day quarantine period for the evidence of transmission from their associated index cases (**Fig. 1A**). We found that 48.2% (94/195) of these index cases had at least one contact who became SARS-CoV-2-positive, whereas 51.8% of the index cases (n=101) were non-spreader with no contacts who subsequently tested positive.

**Figure 1.**
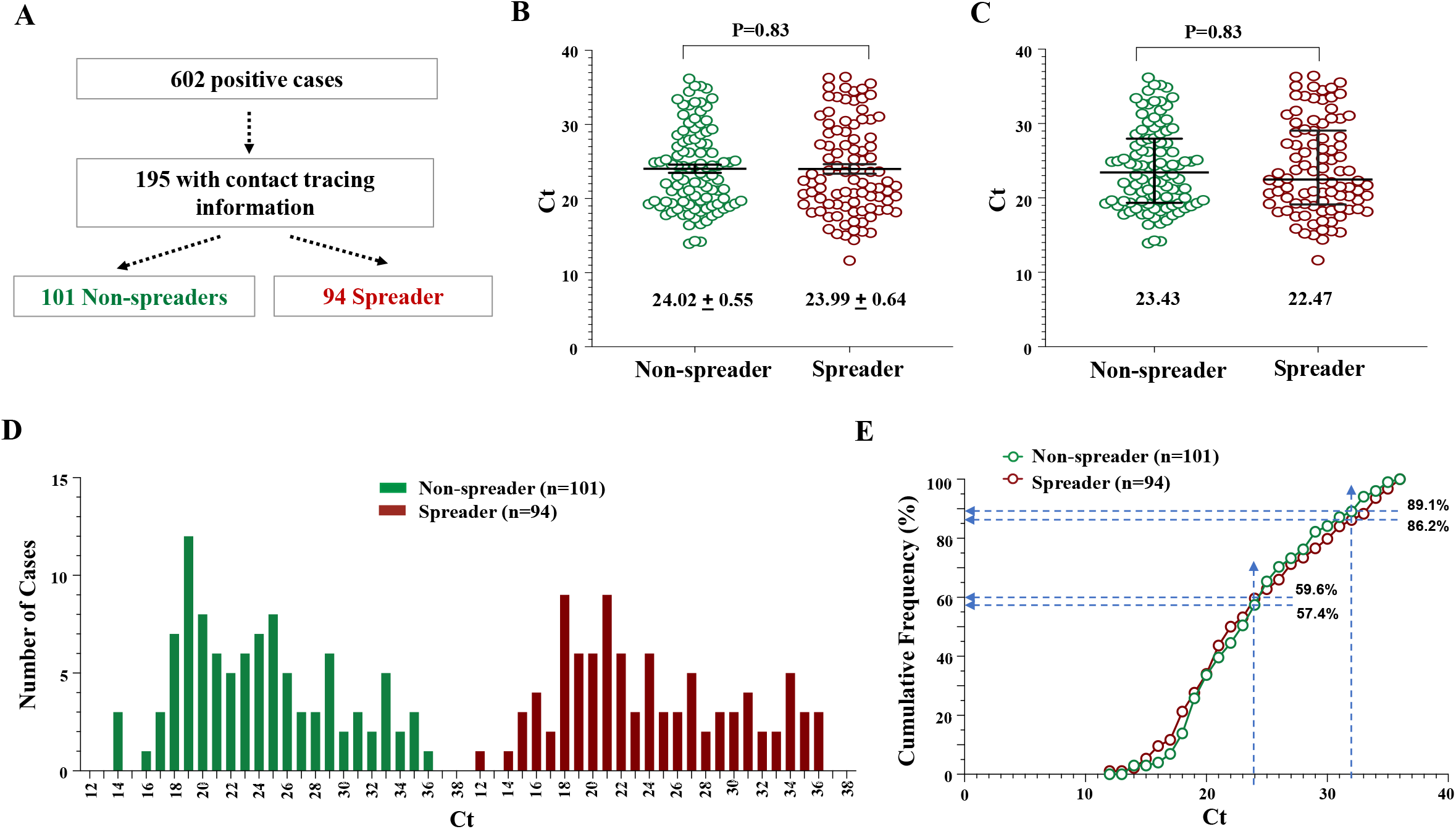
The Ct values of the spreader and the non-spreader are largely overlapped. (**A**). The separation of index cases into the spreader and the non-spreader group. The n of the cases in each population is indicated. (**B-C**). Scattered plots of Ct values expressed as mean value ±SEM (B), or median ± interquartile intervals (C). Ct values of the indicated populations are compared. Mann-Whitney U test. (**D**). The histogram of the distribution of Ct values. (**E**). The cumulative frequency of Ct values. Dashed lines indicate the cumulated percentage of each population at the designated Ct value (24 or 32). At the indicated Ct values and below, there is a higher percentage of spreader cases than non-spreader cases, although the differences are small.

Mean Ct values of the spreader and the non-spreader were nearly identical (**Fig. 1B**), but their median Ct values differed by almost one cycle (**Fig. 1C**), suggesting that more spreaders had a lower Ct value than the non-spreader. However, Ct distributions in these groups were similar with the main peaks around 18-21 (**Fig. 1D**), although the Ct range was slightly broader for the spreader (12-36) than that for the non-spreader (14-36). Cumulative Ct frequencies overlapped between the spreader and the non-spreader with 10.9%, and 13.8% of cases having a Ct value of 32 and higher, respectively (**Fig. 1E**), but the difference was not large enough to discriminate the two groups for practical use.

In a reverse approach, index cases were traced for 481 students undergoing quarantine at one of the three Tulane quarantine sites in September 2020 (**Fig. 2A**), 18% of whom (85/481) became positive during their quarantine period. Index cases for these 481 quarantined individuals were considered spreaders if they were linked to one or more quarantined students with a positive test result and non-spreaders if they were associated only with individuals with negative test results. Spreaders and non-spreaders without Ct reported were excluded from further analysis. We found that mean Ct values of the spreader and the non-spreader groups did not differ (**Fig. 2B**). Taken together, these index case studies suggest that Ct values alone do not predict transmission risk.

**Figure 2.**
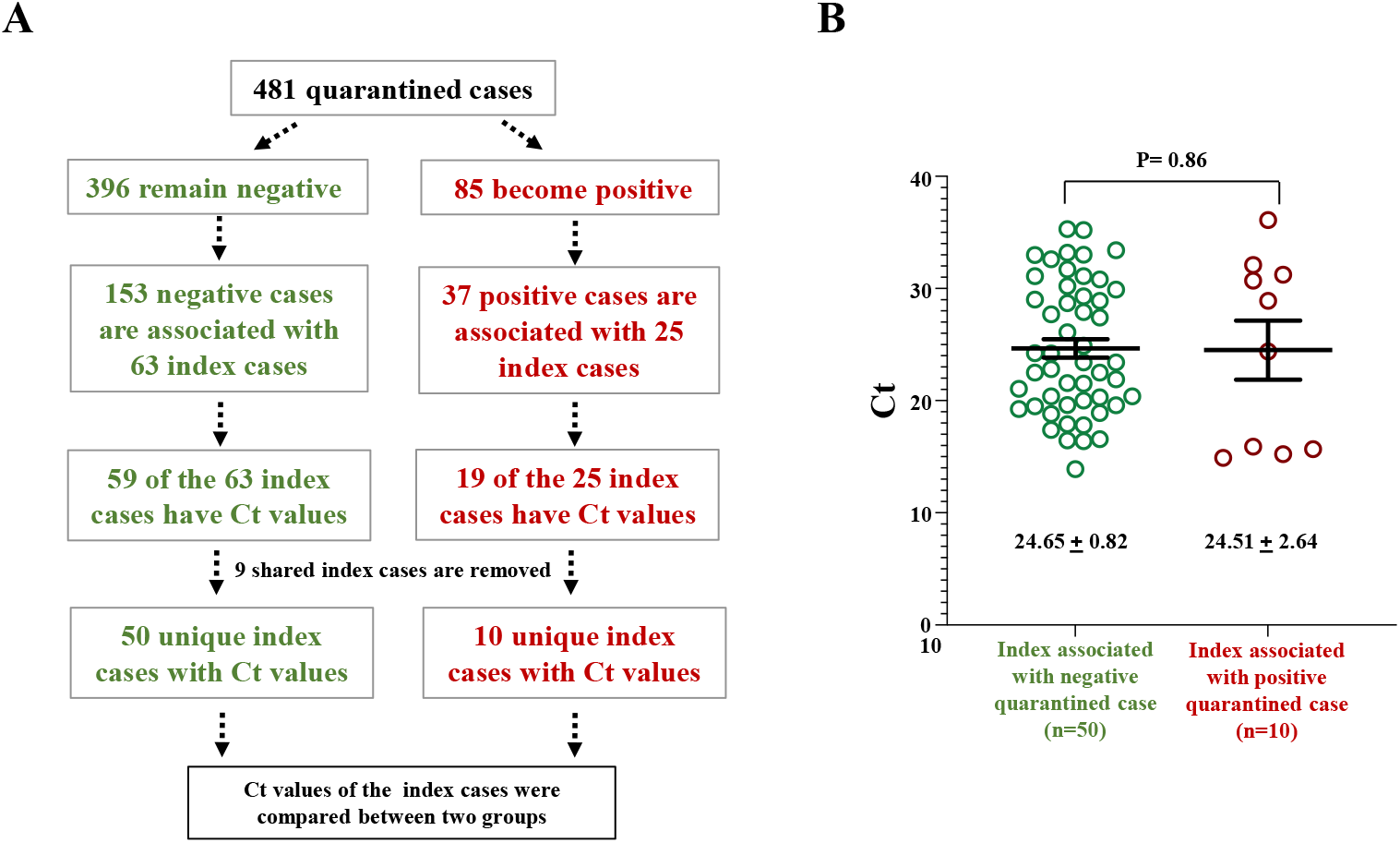
Comparisons of Ct values of index cases tracked from quarantined cases. (**A**). Diagram of the study design. Index cases with Ct values available are tracked down from their contacts in the quarantined unit. (**B**). The Ct values of spreader index cases and non-spreader index cases show a significant overlap. Data shown are mean±SEM. Mann-Whitney U test.

Individuals who are SARS-CoV-2 positive but asymptomatic can still be infectious [12-14], and may exhibit a similar viral load to their symptomatic counterparts [12, 13, 15]. We therefore identified 375 positive cases who were evaluated for COVID-19 symptoms at testing (**Fig. 3A**) to assess the relationship between symptom presentation and Ct values. We found that the mean and median Ct values were significantly lower in symptomatic than those in asymptomatic cases (**Fig. 3B-C**), which was also reflected by the difference in the Ct range of these groups (12-36 versus 14-37; **Fig. 3D**). Although both groups exhibited Ct peaks around Ct 19-22, there was a noticeable rightward shift in the cumulative Ct frequency in the asymptomatic versus symptomatic population, indicative of reduced viral load in the asymptomatic group (**Fig. 3E**). In comparison, other studies with cohorts differing in location and in constituents, including a large study involving senior citizens from nursing houses and assisted living facilities in Massachusetts, found that Ct values did not differ significantly between the symptomatic and the asymptomatic individuals; but observed a faster virus clearance, as measured by Ct value, in the asymptomatic cases than in the symptomatic cases [13, 15]. These and our studies thus suggest that infections with a higher viral load may more likely lead to symptom development, or that symptomatic persons tend to have higher viral loads or to maintain their viral loads for a longer time.

**Figure 3.**
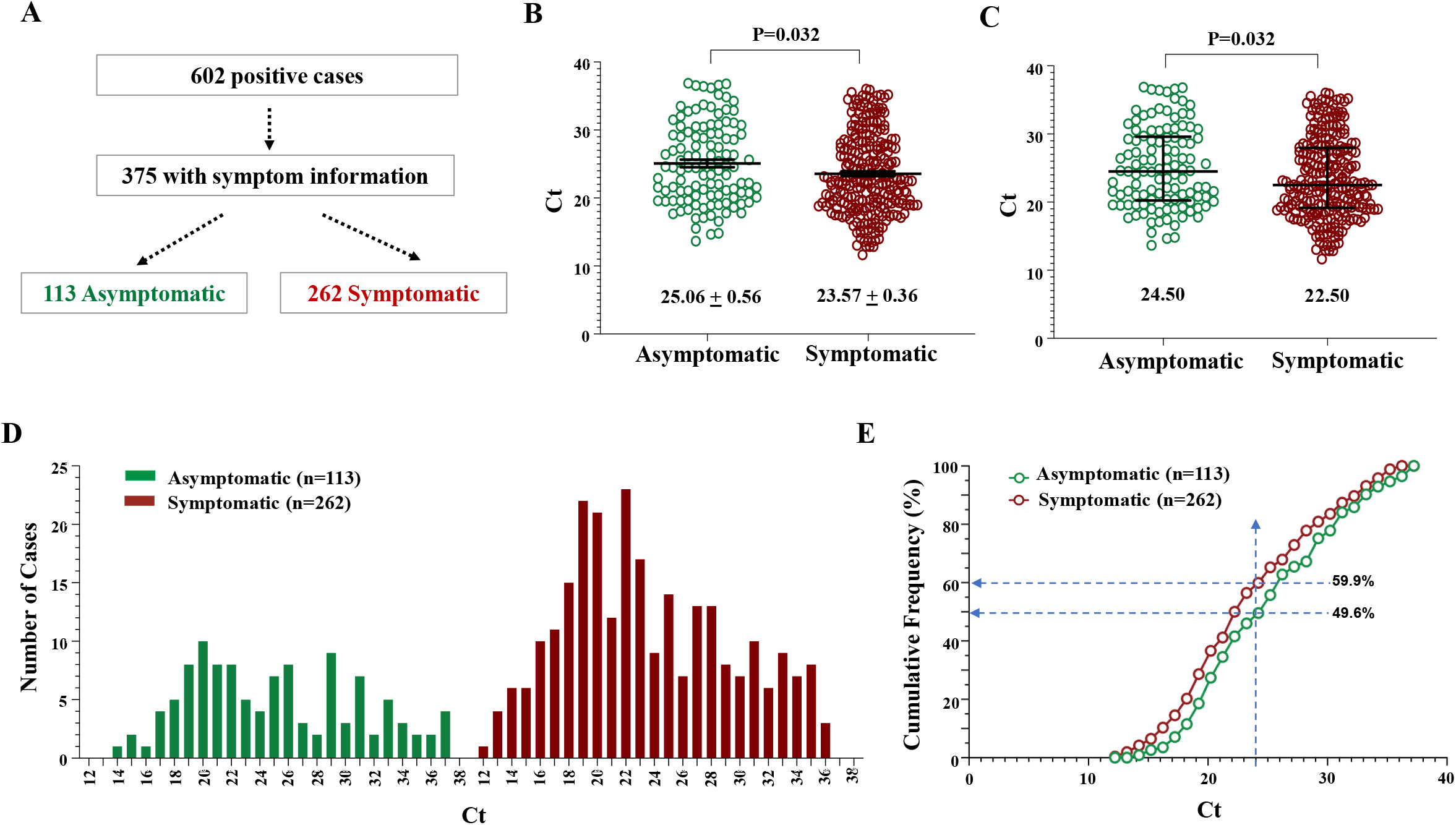
The Ct values of the spreader and the non-spreader are largely overlapped. (**A**). The separation of positive cases into the symptomatic and the non-symptomatic groups. The n of the cases in each population is indicated. (**B-C**). Scattered plots of Ct values expressed as mean value ±SEM (B), or median ± interquartile intervals (C). Ct values of the indicated populations are compared. Mann-Whitney U test. (**D**). The histogram of the distribution of Ct values. (**E**). The cumulative frequency of Ct values. Dashed lines indicate the cumulated percentage of each population at the Ct value of 24. At this Ct value and below, there is a higher percentage of symptomatic cases (59.9%) than asymptomatic cases (49.6%).

All 195 index cases with contact tracing information had information recorded regarding symptoms. We thus further divided the spread group and the non-spreader group based on symptom presentation (**Fig. 4A**). We found that the symptomatic spreader had the lowest mean and median Ct values, differing by 2 cycles for the mean and 3.5 cycle for the median when compared with the asymptomatic non-spreader, which had the highest mean and median Ct values (**Fig. 4B-C**). The Ct distribution indicated that the symptomatic groups (spreader and the non-spreader) and the spreader groups (with or without symptoms) tended to have more individuals with lower Ct values (<24) (**Fig. 4D-E**). This finding suggests that SARS-CoV-2 spreaders tend to have more virus and are more likely symptomatic.

**Figure 4.**
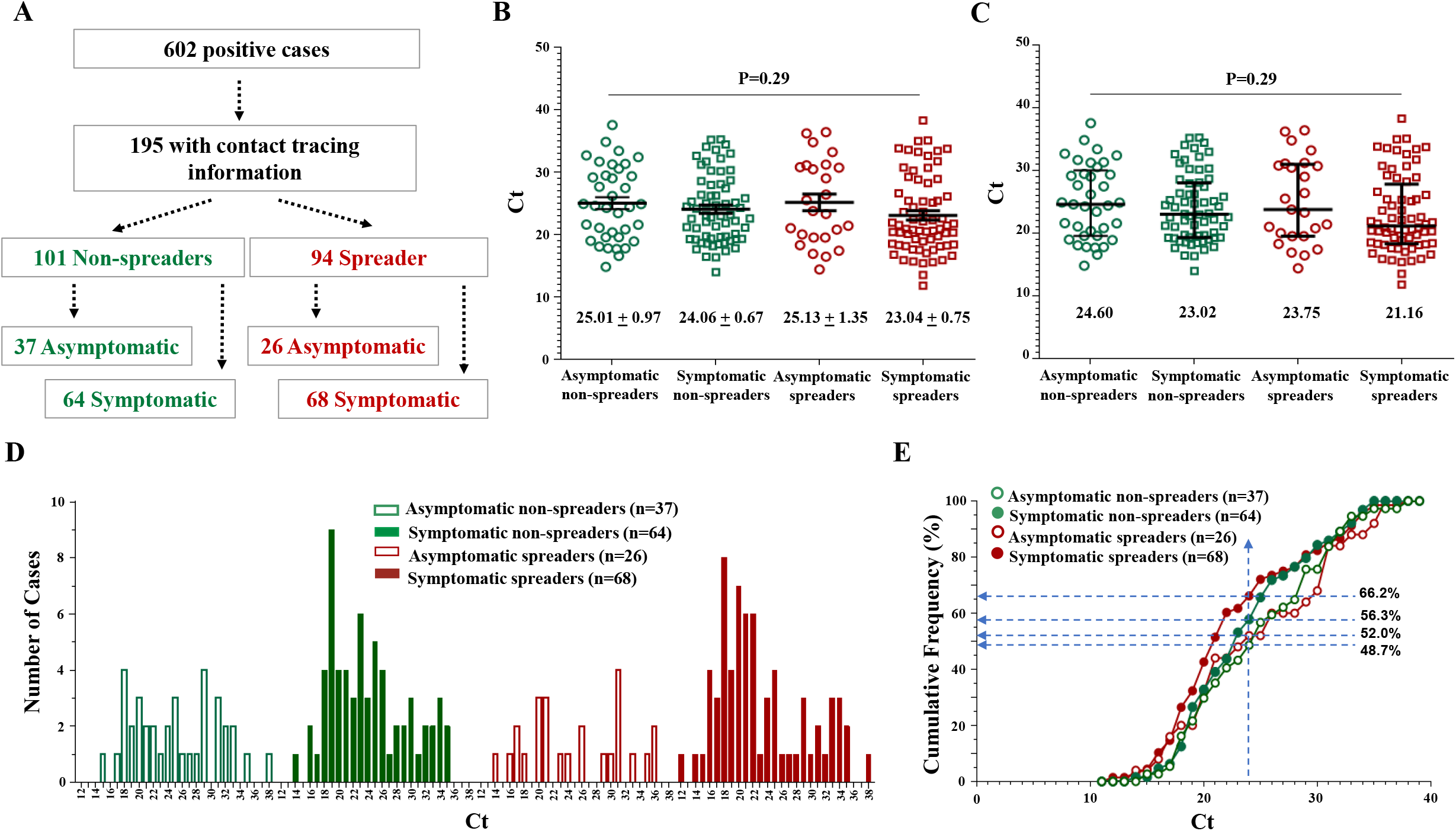
The Ct values of the spreader and the non-spreader are largely overlapped. (**A**). The separation of positive cases into the spreader and non-spreader with or without symptoms. The n of the cases in each population is indicated. (**B-C**). Scattered plots of Ct values expressed as mean value ±SEM (B), or median ± interquartile intervals (C). Ct values of the indicated populations are compared. One-way ANOVA. (**D**). The histogram of the distribution of Ct values. (**E**). The cumulative frequency of Ct values. Dashed lines indicate the cumulated percentage of each population at the Ct value of 24. At this Ct value and below, there is a higher percentage of symptomatic spreader cases (66.2%) than asymptomatic non-spreader cases (48.7%). The percentage of cases of the other groups are between the two.

## Discussion

The present study for the first time compared the Ct values between the spreader and the non-spreader of SARS-CoV2 infected persons in a college student population. We have found that while the mean Ct values of the spreader, particularly the symptomatic spreader, is lower than the non-spreader, there are significant overlaps among individuals, whether they are spreaders or non-spreaders. It is thus practically not feasible to predict who would be spreaders or not based on the viral load as detected from their nasal swabs.

Ct values are not reported in current public health practice despite that they may be informative of viral burden. Our study supports this practice and indicates that, due to broad spread and overlap in Ct values across the spectrum of symptom presentation and transmissibility, Ct value reporting at the individual level, such as by setting a cut-off value at 32 [5-10], would provide little diagnostic value for differential case management. At the population level, Ct values may be useful, particularly in association with the symptomatic presentation, to indicate the likelihood of transmission. It may thus have epidemiological or surveillance values.

Detection of SARS-CoV-2 may need to be both sensitive and rapid, which may not always be feasible for all methods. Rapid but less sensitive method should be used more frequently in order to catch individuals whose virus level may be elevating over the course and thus presumably become more infectious. However, our results suggest that individual with low viral load could still be infectious. Thus sensitive and robust SARS-CoV-2 diagnostic testing method is needed to effectively control the viral transmission by maximizing the ability to identify and quarantine those infected with a low level of virus and.

Although limited by its retrospective nature, this study likely benefits from less interference from host and environmental factors on viral transmission, since the college student population is generally in good health with few underlying susceptibilities, with most individuals living and interacting in a shared and relatively confined social environmental (*i*.*e*., campus). Transmissibility is not only affected by the viral load of the spreader, and the environment where transmission takes place, but also affected by factors that underline the susceptibility of the population, such as the age, gender, and the basic health conditions. From this aspect, it is interesting to note that while male and female students had nearly the same proportion of the infected, consistent with a meta-analysis of 90 reports [11] the male students did have a higher infection rate (10.65%) than the female students (6.56 %) in this cohort. The gender disparity of COVID-19 has been well recognized in terms of the severity of the disease with the male being more likely to develop severe conditions [11]. It has yet to be determined how the gender makes the difference in the COVID-19 spread and development.

In summary, this study has determined that Ct values of the spreader may be lower at the population level than the non-spreader, but the large overlap in the values at the individual level prevents their use as a differential tool to guide isolation and quarantine practice. On the other hand, a sensitive and robust diagnostic method is necessary to restrict viral transmission from those carrying a low level of virus.

## Data Availability

Data available within the article or its supplementary materials

## Acknowledgement

This study is supported by Tulane University.

## Contribution of the authors

Concept and design: D. Tian, R. Garry, X-M. Yin, P. Delafontaine, L. Hamm

Data acquisition and analysis: D. Tian, E. Kriner, J. DeVoto, N. Okami

Drafting of the manuscript: X.-M. Yin, D. Tian, Z. Lin,

Editing: T. Hu, R. Garry, P. Delafontaine

Administrative and operation support: J. Esneault, M. Livaccari, M. Lampp, S. Tim, P. Norton IT support: J. Tran, N. Tran, N. Wang

